# A transdiagnostic evaluation of adult temperamental profiles and their interactions with childhood adversities across major psychiatric disorders

**DOI:** 10.1101/2025.11.12.25340048

**Authors:** Pavithra Dayalamurthy, Srinivas Balachander, Sai Priya Lakkireddy, Mahashweta Bhattacharya, Mino Susan Joseph, Pramod Kumar, Anand Jose Kannampuzha, Sreenivasulu Mallappagari, Shruthi Narayana, Alen Chandy Alexander, Moorthy Muthukumaran, Joan C Puzhakkal, Navya Spurthi Thatikonda, Sweta Sheth, Furkhan Ali, Sowmya Selvaraj, Vanteemar S Sreeraj, Bharath Holla, Ganesan Venkatasubramanian, John P. John, Biju Viswanath, cVEDA Consortium, ADBS-CBM Consortium, Vivek Benegal, YC Janardhan Reddy, Sanjeev Jain

**Author notes:** Equal contribution as first author (Pavithra Dayalamurthy, Srinivas Balachander). **Correspondence:** Srinivas Balachander, Assistant Professor, Department of Psychiatry, National Institute of Mental Health and Neuro Sciences (NIMHANS), Bangalore, Karnataka, India 560029; Biju Viswanath, Additional Professor, Department of Psychiatry, NIMHANS, Bangalore, Karnataka, India 560029. **ADBS-CBM Consortium:** Biju Viswanath, Venkatasubramanian Ganesan, John P John, Meera Purushottam, Reeteka Sud, Bharath Holla, Jayant Mahadevan, Srinivas Balachander, Sreeraj VS, Vijay Kumar K G, Suhas Ganesh, Suhas Satish, Preethi V Reddy, Vijaykumar S Harbishettar, Lekhansh Shukla, Pradip Paul, Bhagyalakshmi M.S, Palanimuthu T Sivakumar, Arun Kandasamy, Muralidharan Kesavan, Urvakhsh Meherwan Mehta, Ashitha S.N.M, Bhupesh Mehta, Thennarasu Kandavel, B Binu Kumar, Jitender Saini, A Shyamsundar, Gautam Arunachal Udipi, Himani Kashyap, Anish V Cherian, K S Meena, Latha K, Jagadisha Thirthalli, Prabha S Chandra, Pratima Murthy, Upinder S Bhalla, Mathew Varghese, Sanjeev Jain, Vivek Benegal, Padinjat Raghu, Janardhan Y C Reddy Department of Psychiatry, NIMHANS, Bangalore; Department of Biostatistics, NIMHANS, Bangalore; Department of Neuroimaging and Interventional Radiology, NIMHANS, Bangalore; Department of Biophysics, NIMHANS, Bangalore; National Centre for Biological Sciences (NCBS), Bangalore; Institute of Stem Cell and Regenerative Medicine (InStem), Bangalore. **cVEDA Consortium:** Eesha Sharma, Nilakshi Vaidya, Sunita Simon Kurpad, Rebecca Kuriyan-Raj, Debasish Basu, Subodh Bhagyalakshmi Nanjayya, Kalyanaraman Kumaran, Ghattu Krishnaveni, Amit Chakrabarti, Rajkumar Lenin Singh, Roshan Lourembam Singh, Kartik Kalayaram, Rose Dawn Bharath, Dimitri Papadopoulos Orfanos, Mathew Hickman, Mireille Toledano, Yuning Zhang, Gunter Schumann Department of Child and Adolescent Psychiatry, NIMHANS, Bangalore; Centre for Population Neuroscience and Precision Medicine (PONS), Institute of Psychiatry, Psychology & Neuroscience, SGDP Centre, King’s College London, London, UK; Department of Psychiatry, St John’s Medical College and Research Institute; Department of Psychiatry, Post Graduate Institute of Medical Education and Research (PGIMER), Chandigarh; Epidemiology Research Unit, CSI Holdsworth Memorial Hospital, Mysore; Foundation for Research and Advocacy in Mental Health, Mysore; ICMR-Centre for Ageing and Mental Health (I-CAM), Division of Non-Communicable Diseases (NCD), Indian Council of Medical Research (ICMR), Kolkata; Regional Institute of Medical Sciences, Imphal; Rishi Valley School, Madanapalle; Department of Neuroimaging and Interventional Radiology, NIMHANS, Bangalore; NeuroSpin, CEA, Université Paris-Saclay, Gif-sur-Yvette, France; Centre for Population Neuroscience and Precision Medicine, MRC Social, Genetic, Developmental Psychiatry Centre, Institute of Psychology, Psychiatry & Neuroscience, King’s College London, London, UK; MRC Centre for Environment and Health, School of Public Health, Imperial College London, London, UK; Centre for Population Neuroscience and Precision Medicine, Charité Mental Health, Department of Psychiatry and Psychotherapy, Charité Universitätsmedizin, Berlin, Germany.

## Abstract

**Background:** Temperamental profiles and adverse childhood experiences (ACEs) have been reported in most major psychiatric disorders. However, the nature of this relationship is complex and under-studied.

**Methods:** A total of 1162 individuals (N=532 affected; N=457 unaffected first-degree relatives (FDRs) and N=173 healthy controls) were included. Affected individuals and their FDRs were from 393 multiplex families of schizophrenia, bipolar disorder, obsessive-compulsive disorder, or alcohol use disorder. They were assessed using the self-reported Adult Temperament Questionnaire (ATQ), and the ACEs International Questionnaire. We performed frequentist and Bayesian mixed-effects regression models, to look for associations between the ATQ domains, ACEs score and their interaction with each diagnosis/familial risk. External validity of our final model was tested to predict diagnosis in an independent cohort. Receiver operator characteristic plots and the area under the curve (AUC) were used to evaluate its prediction accuracy.

**Results:** All temperamental domains: negative affectivity (NA), extraversion (EV), effortful control (EC) and orienting sensitivity (OS) had significant associations in a unique configuration within each of the four diagnoses and their familial risk. ACEs, and its interactions were significantly associated with alcohol use disorder only. In the external validation dataset, the final model was found to successfully predict AUD (AUC=0.82), BD (AUC=0.75) and OCD (AUC=0.74), but not SCZ (AUC=0.53).

**Conclusions:** These findings provide unique insights into the complexities of temperament in major psychiatric disorders, and emphasize the importance of evaluating temperament in conjunction with childhood adversity to characterize risk.

Graphical Abstract (Study Methodology):

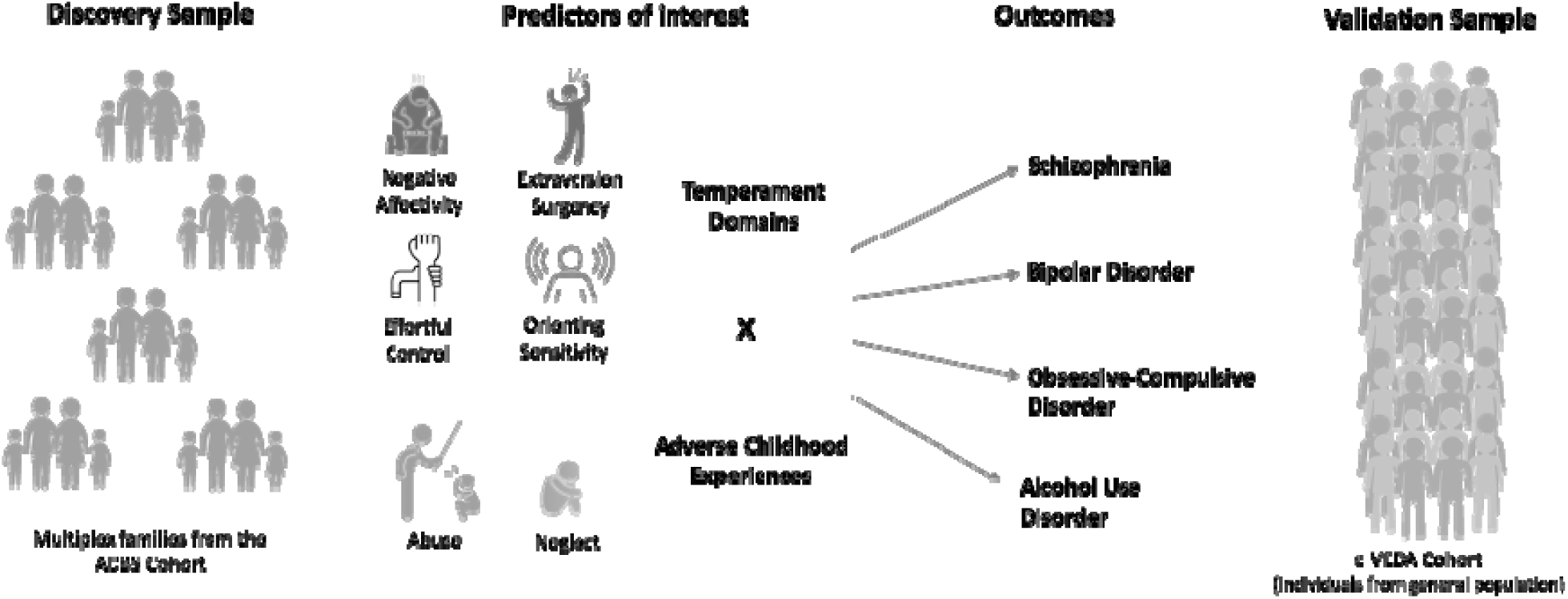

## Introduction

There is an increasing appreciation from neurobiological, genetic and clinical studies that a developmental perspective is crucial to understand psychiatric disorders (Cicchetti & Curtis, 2015; Rutter, 1988). Recognition of developmental markers may be helpful in early identification, targeted treatments and even prevention of psychiatric syndromes (Thapar & Riglin, 2020). One such marker, frequently investigated and linked to psychiatric disorders is temperament (Rettew & McKee, 2005). Temperament refers to the constellation of inborn traits (individual disposition of particular patterns of emotional reactions, mood shifts and sensitivity to stimulations), that determine the individual’s unique behavioral style in experiencing and reacting to the world (Goldsmith et al., 1987). Dimensions of temperament are recognizable early in life, and understood to be relatively stable throughout life (Bornstein et al., 2019; Kopala-Sibley et al., 2018).

Temperament is traditionally evaluated in children/adolescents by behavioral observations or parental reports. Over the last three decades, methods to measure temperament optimally in adults have undergone significant developments. Most contemporary studies have evaluated temperament in adults with the Temperament-Character Inventory (TCI) (Cloninger et al., 1994) or more recently, the Rothbart Adult Temperament questionnaire (ATQ) (Evans & Rothbart, 2007). Both these instruments conceptualize temperament as having four “core” dimensions: 1) the novelty-seeking/reward dependence (TCI) or extraversion/surgency (ATQ) dimension (the ‘behavioral activation/approach’ system); 2) negative affectivity (ATQ) or harm avoidance (TCI) (the ‘behavioral inhibition’ system), 3) effortful control (ATQ) or persistence (TCI) (the ‘executive’ system) that helps regulate the first two dimensions; and 4) orienting sensitivity (ATQ) (included both affective and general perceptual sensitivity). The self-rated ATQ is briefer (77 items) compared to the TCI (240 items), and has been shown to be psychometrically robust and translated into various languages (Laverdière et al., 2010; Wiltink et al., 2006).

Longitudinal studies have shown that early temperamental differences exert a pervasive influence on subsequent life-course development, personality structure, interpersonal relations, and psychopathology (Caspi, 2000). While the early research on temperament, focused on children and adolescents and found associations with several childhood psychiatric disorders (childhood depression, anxiety disorders, attention-deficit hyperactivity disorder, and other disruptive behavior disorders), recent studies have begun looking at the relationship between childhood temperament and subsequent adult-onset psychiatric disorders such as schizophrenia (SCZ), bipolar disorder (BD), obsessive-compulsive disorder (OCD) and alcohol use disorders (AUD) (Miettunen & Raevuori, 2012).

While the common strategy for identifying risk factors is to identify variables associated with heightened risk for specific psychiatric disorders, researchers are increasingly identifying transdiagnostic risk factors that are shared across disorders (Sauer-Zavala et al., 2017). Meta-analyses of temperament in various psychiatric disorders (Miettunen, 2011; Miettunen & Raevuori, 2012; Ohi et al., 2012) have found several temperamental traits to be transdiagnostic. Inhibited temperament, for example, characterized by fearful or avoidant response to novelty, long thought to be a vulnerability for the development of anxiety disorders (Biederman et al., 2001), has now been demonstrated to confer a broader risk across many psychiatric disorders, being similarly elevated in patients with SCZ, BD, and major depressive disorder (Feola et al., 2020). Higher harm avoidance has been reported in SCZ (Sim et al., 2012; Smith et al., 2008) and OCD samples (Bey et al., 2017; Ettelt et al., 2008), and also in their first-degree relatives. High reward dependence, lower persistence have been reported in AUD (Wills & Dishion, 2004) and BD samples (Almeida et al., 2011; Porcelli et al., 2017). Lower ‘effortful control’ (or higher impulsivity) is another trait reported commonly in OCD (Coles et al., 2006), AUD (Dick et al., 2010), as well as BD (Strakowski et al., 2010). In essence, the transdiagnostic nature of these vulnerability factors may help explain the co-occurrence of symptoms, comorbidities, or familial loading across psychiatric disorders (Fusar-Poli et al., 2019; Sreeraj et al., 2021).

Temperament is shown to be heritable, and across populations there is a strong tendency for closely related individuals to measure similarly (Cloninger et al., 2019). Similar temperamental patterns that precede or confer vulnerability for subsequent adult onset disorders, may also be seen in unaffected family members. Though historically recognized (Bleuler, 1974), studies on unaffected “high-risk” individuals from families of these disorders are far fewer. Studying both (affected and unaffected “at-risk” individuals) would provide valuable insights into etiological processes or the type of relationship shared between them. Although temperamental traits have not always been recognized as typical “endophenotypes”, they could be considered as such, because they are normally distributed in the general population, are more homogenous than the clinical phenotype of psychiatric disorders, are relatively stable, appear early in life, and show high heritability (Caspi et al., 2005; McCrae et al., 2000). The nature of the relationship between temperament and psychiatric disorders is thus complex, and multiple hypothetical pathways have been invoked (Horan et al., 2008; John et al., 2010).

Children’s temperaments contribute significantly to their interactions with their environments. The original temperament studies (Thomas & Chess, 1977) argued that adaptive childhood outcomes occurred when the temperamental qualities of a child were “congruent with environmental characteristics suited for that temperament type”, a concept referred to as “goodness of fit”. Maladaptive outcomes were considered to be a consequence of the mismatch between the child’s proclivities and their environment, rather than either of these factors in isolation. In this context, one would need to consider the role of environmental risk factors, such as adverse childhood experiences (ACEs). Among the ACEs, childhood maltreatment (which includes neglect, physical and sexual abuse) is found to be associated with most major psychiatric disorders, and is again considered a transdiagnostic risk factor (McLaughlin et al., 2020). Few studies have evaluated the complex interplay between temperamental and environmental factors in major psychiatric disorders.

Using large data from an ongoing longitudinal study in India, we thus aimed to: i) explore the overlap of temperamental profiles/patterns in multiplex families with major psychiatric disorders, identify if they are shared within families, between subjects with psychiatric illness and their unaffected first-degree relatives (FDRs); ii) identify how specific temperamental traits are associated with either the diagnosis, severity, and the familial risk of different major psychiatric disorders: schizophrenia (SCZ), bipolar disorder (BD), obsessive-compulsive disorder (OCD), alcohol use disorder (AUD); iii) evaluate interactions between these temperamental domains and ACEs. Finally, we aimed to test the external validity of our findings in a large, independent, population-based sample. We hypothesized that: i) temperamental traits would be shared within families; ii) each psychiatric disorder would have a unique temperamental profile, which would correlate with illness severity and also be present in unaffected FDRs with its familial risk; iii) these temperamental factors would be significantly moderated by ACEs; iv) our results would be externally valid.

## Methods

### Study sample

We used data from the Accelerator program for Discovery in Brain Disorders using Stem cells (ADBS) project, an ongoing longitudinal study, set up to study the developmental trajectories and basic biology of major psychiatric disorders (Viswanath et al., 2018). The study sample comprises a cohort of psychiatrically ill participants from multiplex families, their unaffected FDRs and matched healthy participants from unaffected families. ‘Multiplex families’ are those wherein multiple members (at least two affected FDRs in a nuclear family) are diagnosed to have major psychiatric disorders (SCZ, BD, OCD, AUD and Alzheimer’s dementia). Familial loading was verified by applying the Family Interview for Genetic Studies (FIGS) (Maxwell, 1996), along with a pedigree derived by interviewing at least three members of the family. The psychiatric diagnosis (or lack thereof in unaffected FDRs) was corroborated using the Mini International Neuropsychiatric Interview (MINI) (Sheehan et al., 1998) by direct interviews of all participants. The clinical characteristics in the sample such as diagnosis, comorbidity, intra-familial co-occurence of different syndromes, have been described in an earlier paper (Sreeraj et al., 2021). The healthy control group consisted of volunteers aged between 16 to 60 years who were screened for the absence of any current/lifetime psychiatric diagnosis in self as well as in any FDR as per the MINI and the FIGS respectively. Those with Alzheimer’s dementia were not included for this analysis. The project was reviewed and approved by the Institutional Ethics Review Board and written informed consent was obtained from all participants.

### Assessments

#### Adult Temperament Questionnaire

We used the short form of the self-report ATQ (Evans & Rothbart, 2007), which has 77 items. Each item was based on the 7-point Likert scale ranging from “extremely untrue” to “extremely true”. The scale was adapted in five different Indian languages (Kannada, Tamil, Telugu, Malayalam & Hindi), using standard procedures for translation and back-translation (World Health Organization, 2013). The ATQ dimensions have been validated (Rothbart et al., 2000) to group into four main factors: Negative Affectivity (NA), Extraversion/Surgency (EV), Effortful Control (EC), and Orienting Sensitivity (OS); as well as smaller sub-factors. **Supplementary Table 1** shows the reliability measures (internal consistency) within the various factors and sub-domains of the scale for each of the translations. We included data from participants who completed at least 80% (i.e, 61 of the 77 questions) in the ATQ.

#### Adverse Childhood Experiences

The WHO ACE-IQ questionnaire (World Health Organization, 2018), consists of 31 questions across 13 subdomains: physical abuse; emotional abuse; contact sexual abuse; alcohol and/or drug abuser in the household; incarcerated household member; household member with a psychiatric condition or suicidality; household member treated violently; one or no parents, parental separation or divorce; emotional neglect; physical neglect; bullying; community violence; and collective violence. The total ACE severity score was calculated by adding the total number of subdomains where adversity was reported to have occurred above a predefined frequency threshold (aka “Frequency Score”). This threshold differs across the subdomains of ACE reported (e.g. contact sexual abuse only requires being touched sexually once, but emotional abuse requires being screamed at many times).

#### Clinical Global Impression – Severity

Cross-sectional illness severity was measured using this widely-used tool (Guy, 1976). The overall severity is rated on a likert scale ranging from 1 (not at all ill) to 7 (among the most extremely ill patients).

All clinician-administered assessments (MINI, FIGS, ACE-IQ, CGI-S) were performed by trained clinical psychologists, psychiatric social workers, or psychiatrists, all of whom underwent regular training in the use of the instruments.

### Validation Dataset

We used data from the Consortium for Vulnerability to Externalizing Disorders and Addiction (c-VEDA) study (Sharma et al., 2020; Zhang et al., 2020). This is a large (N=9010) neurodevelopmental cohort of children, adolescents, and young adults sampled from 5 sites across India. Subsetting only the adults (age range 18-23 years) within this dataset, in which the same assessments (MINI, ATQ and ACE-IQ) were available, yielded an effective sample of N=2485 from this cohort.

### Statistical Analysis

Descriptives of the sociodemographic details and ATQ domain scores in the sample were done using mean/standard deviations and n/percentage. The affected probands or FDRs groups for each diagnosis were not mutually exclusive (i.e., an individual proband, or FDR could have multiple comorbid diagnoses, or be “at-risk” for multiple diagnoses), no statistical analyses were done to compare the diagnostic/FDRs groups with each other. Normality was checked for all the variables by examining histograms and Q-Q plots. Variables that were found to have a non-normal distribution were transformed using automated algorithms in the “bestNormalize” package in R, version 1.8.2 (Peterson, 2021). Missing data from the ATQ was imputed using a predictive mean matching algorithm in the package “mice” in R, version 3.14.0.

Regression models with both frequentist as well as Bayesian approaches were used to examine relationships between each temperamental domain, ACEs, and each disorder along with its familial risk. First, we used linear mixed-effects regression models to predict each of the ATQ domains, by diagnoses or familial risks. Family ID was used as a random-effects predictor in all models, and intra-class correlation coefficients (ICCs) were derived to estimate the degree of intra-familial sharing for each temperamental domain. Two separate sets of regression models were used, one combining the affected probands and healthy controls, and another including only the unaffected FDRs and healthy controls. Dummy-coded categorical diagnosis or familial risk variables for each disorder were used as predictor variables in the regression models. Age, gender and education were included as covariates. In order to account for familial loading of other disorders in many of the affected participants, we performed a sensitivity analysis, in which we regressed out the effects of familial risk from the temperamental domains, and used the residuals as outcome variables. The lme4 package (Bates et al., 2015) was used for running these regression models. As there were 4 ATQ domains, we used a Bonferonni-corrected p-value threshold of 0.05/4 = 0.0125.

Bayesian regression models using the brms package (Bürkner, 2017, 2018), were used to run a complex multinomial logistic regression model, with each dummy-coded diagnosis as the dependent variables, and the temperamental domains (regressed out for age, gender and education), ACEs total score and the interaction between ACEs x each temperamental domain as predictors. Default priors were used as chosen by the program, as informed priors were not feasible, given the non-random manner in which our sample was obtained, and the complexity of our model with many parameters and variance components. The Markov Chain Monte Carlo (MCMC) sampler was run in 4 chains of 10,000 iterations (with a warm-up of 1000 iterations) and convergence was checked using the R-hat (*R̂*) statistic, along with the bulk and tail Effective Sample Sizes (ESS). Model performance was evaluated using the leave-one-out (LOO) cross-validation procedure. We calculated the Bayes factor between the model with temperamental domains x ACE interaction terms (H_1_) over the model with only the main effects of temperament and ACEs individually (H_0_). We report and interpret the mean posterior probabilities, and their 95% credible intervals of the main study model. The reporting of this analysis and results is in accordance with the recently published Bayesian Analysis Reporting Guidelines (BARG) (Kruschke, 2021).

The external validation was performed by using the multinomial logistic regression model to predict the same diagnoses in the c-VEDA dataset. For each diagnosis, the true positivity and false positivity rates were obtained, using which receiver-operator-characteristic (ROC) curves were plotted. The area under the curve (AUC) was used as the main index of accuracy of the prediction model We interpret the AUC values as follows: 0.5 = “chance” predictions, 0.5 - 0.7 = poor prediction; 0.7 - 0.8 = acceptable prediction; 0.8-0.9 = excellent prediction; >0.9 = outstanding prediction (Hosmer & Lemeshow, 2000)

## Results

A total of 1162 participants (affected, unaffected ‘at-risk’ FDRs & healthy controls) had the required clinical, ATQ and ACE-IQ data available in the first wave of the ADBS project. **Table 1** shows the clinical and demographic characteristics of the sample included in this study. Out of the total sample, n=532 were affected with SCZ, BD, SUD, OCD or any combination of the four as comorbid conditions; and n=457 were unaffected FDRs. These participants were from a total of 393 multiplex families, while the remaining n=173 were healthy controls. **Figure 1** depicts the actual number of participants with each diagnosis, comorbidity, and familial risk profile. As seen, there were significant overlaps in the diagnostic and familial risk profiles. That is, several unaffected participants had different diagnoses in one or multiple FDRs, thus having multiplex risk profiles. Notable differences in the demographic profile were as follows: lower age and greater educational attainment among healthy controls, a higher proportion of males in the AUD-affected group. These three variables (age, gender and education) were thus used as covariates for subsequent analysis. **Supplementary Table 2** shows the raw scores of the ATQ domain scores within each diagnostic and risk profile group.

**Figure 1.**
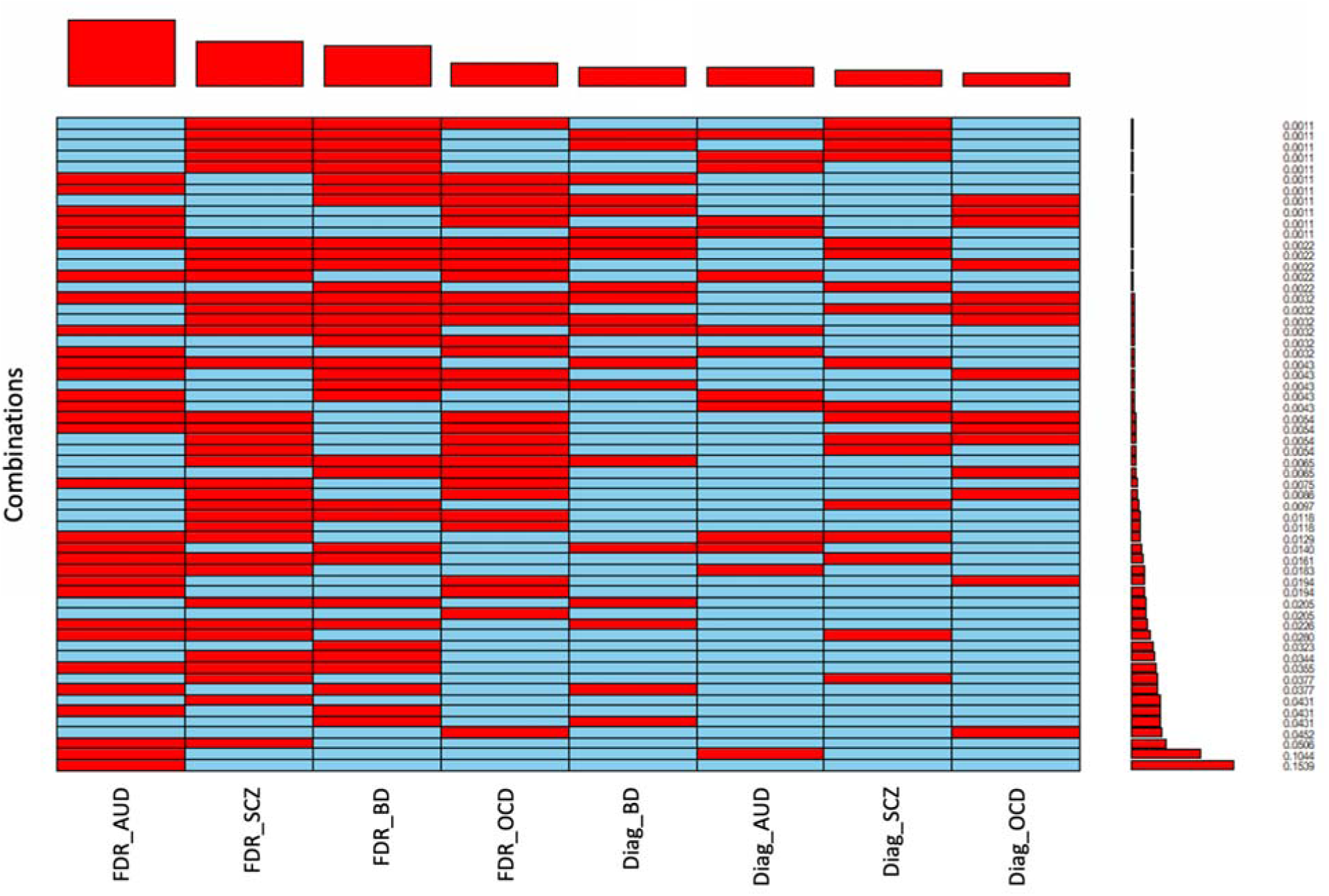
Diagnostic and familial risk profiles in the total sample (N=1162) Each **row** represents a particular combination of diagnosis (Diag_) or familial risk (FDR_) of the disorders under study – alcohol use disorder (AUD), schizophrenia (SCZ), Bipolar disorder (BD) and obsessive-compulsive disorder (OCD). Red indicates presence of that diagnosis among any first-degree relative of the subject, while blue indicates its absence. The bars and numbers on the right indicate the proportion of that combination within the total group, while the bars on top indicate the overall frequency of the diagnosis/familial risk.

**Table 1.** Demographic characteristics of the full sample (Total N=1162; Affected=523; Unaffected FDRs=457; Healthy Controls=173)

|  | Schizophrenia |  | Bipolar Disorder |  | Obsessive-Compulsive Disorder |  | Alcohol Use Disorder |  | Population Healthy Controls |
| --- | --- | --- | --- | --- | --- | --- | --- | --- | --- |
|  | Affected | FDR | Affected | FDR | Affected | FDR | Affected | FDR |  |
| n | 235 | 188 | 228 | 156 | 142 | 75 | 324 | 306 | 173 |
| Age in years | 38.44 (12.12) | 40.07 (14.15) | 37.98 (12.05) | 38.03 (14.83) | 36.71 (13.15) | 39.39 (13.34) | 38.88 (11.66) | 38.15 (14.10) | 39.80 (16.09) |
| Gender (Male) | 131 (55.7%) | 90 (47.9%) | 125 (54.8%) | 83 (53.2%) | 89 (62.7%) | 44 (58.7%) | 237 (73.1%) | 126 (41.2%) | 92 (53.2%) |
| Years of Education | 9.92 (4.77) | 11.02 (4.73) | 10.17 (4.66) | 11.83 (4.04) | 12.37 (3.94) | 12.19 (4.63) | 9.39 (4.64) | 10.40 (4.98) | 13.56 (4.23) |
| Age at Onset | 23.77 (8.87) | - | 22.96 (8.43) | - | 20.78 (9.97) | - | 21.75 (8.35) | - | - |
| Duration of Illness | 14.84 (10.96) | - | 15.43 (10.30) | - | 15.95 (12.07) | - | 17.31 (10.76) | - | - |
| Marital Status (%) |  |  |  |  |  |  |  |  |  |
| Divorced /Separated | 9 (3.8%) | 4 (2.1%) | 7 (3.1%) | 3 (1.9%) | 3 (2.1%) | 1 (1.3%) | 10 (3.1%) | 6 (2.0%) | 3 (1.7%) |
| Married | 117 (49.8%) | 123 (65.4%) | 124 (54.4%) | 94 (60.3%) | 76 (53.5%) | 51 (68.0%) | 210 (64.8%) | 197 (64.4%) | 100 (57.8%) |
| Widowed | 11 (4.7%) | 5 (2.7%) | 10 (4.4%) | 6 (3.8%) | 2 (1.4%) | 1 (1.3%) | 11 (3.4%) | 15 (4.9%) | 9 (5.2%) |
| Single | 98 (41.7%) | 56 (29.8%) | 87 (38.2%) | 53 (34.0%) | 61 (43.0%) | 22 (29.3%) | 93 (28.7%) | 88 (28.8%) | 61 (35.3%) |
| Occupation (%) |  |  |  |  |  |  |  |  |  |
| Unemployed | 53 (22.6%) | 9 (4.8%) | 33 (14.5%) | 8 (5.1%) | 24 (16.9%) | 4 (5.3%) | 51 (15.7%) | 10 (3.3%) | 4 (2.3%) |
| Student | 18 (7.7%) | 19 (10.1%) | 17 (7.5%) | 18 (11.5%) | 21 (14.8%) | 9 (12.0%) | 12 (3.7%) | 45 (14.7%) | 39 (22.5%) |
| Home-maker | 59 (25.1%) | 42 (22.3%) | 58 (25.4%) | 30 (19.2%) | 38 (26.8%) | 17 (22.7%) | 55 (17.0%) | 75 (24.5%) | 23 (13.3%) |
| Employed | 105 (44.7%) | 118 (62.8%) | 120 (52.6%) | 100 (64.1%) | 59 (41.5%) | 45 (60.0%) | 206 (63.6%) | 176 (57.5%) | 107 (61.8%) |
| Language of assessment (%) |  |  |  |  |  |  |  |  |  |
| Hindi | 30<br>(12.8%) | 26<br>(13.8%) | 49<br>(21.5%) | 35<br>(22.4%) | 48<br>(33.8%) | 25<br>(33.3%) | 35<br>(10.8%) | 40<br>(13.1%) | 29<br>(16.8%) |
| English | 11<br>(4.7%) | 16<br>(8.5%) | 17<br>(7.5%) | 7<br>(4.5%) | 15<br>(10.6%) | 7<br>(9.3%) | 18<br>(5.6%) | 20<br>(6.5%) | 10 (5.8%) |
| Kannada | 102<br>(43.4%) | 71<br>(37.8%) | 79<br>(34.6%) | 49<br>(31.4%) | 44<br>(31.0%) | 22<br>(29.3%) | 157<br>(48.5%) | 136<br>(44.4%) | 60<br>(34.7%) |
| Malayalam | 7 (3.0%) | 11<br>(5.9%) | 7<br>(3.1%) | 9<br>(5.8%) | 2 (1.4%) | 3<br>(4.0%) | 8 (2.5%) | 16<br>(5.2%) | 38<br>(22.0%) |
| Tamil | 43<br>(18.3%) | 36<br>(19.1%) | 35<br>(15.4%) | 27<br>(17.3%) | 12<br>(8.5%) | 7<br>(9.3%) | 59<br>(18.2%) | 36<br>(11.8%) | 14 ( 8.1%) |
| Telugu | 42<br>(17.9%) | 28<br>(14.9%) | 41<br>(18.0%) | 29<br>(18.6%) | 21<br>(14.8%) | 11<br>(14.7%) | 47<br>(14.5%) | 58<br>(19.0%) | 22<br>(12.7%) |

**Note:** The sum of the numbers of the affected and FDR groups across the columns will exceed the total number as participants with multiple diagnosis or familial risks have been double-counted here

Significant associations between the covariates (age, gender and education) on the temperamental domains (**Supplementary Table 3**) were as follows : Higher effortful control and lower extraversion with increasing age; lower negative affectivity, effortful control and orienting sensitivity with male gender; and higher orienting sensitivity with education. The ICC values were significant (lower 95% CI > 0) for all temperamental domains, indicating some degree of intrafamilial sharing.

**Figure 2** depicts the effect sizes of the association between each diagnosis or risk profile with the temperamental domains. We found several significant associations for affected probands in various diagnoses, and few associations for the unaffected FDRs (see **Supplementary Table 4** for associations and their p-values). Affected probands with SCZ and BD had a similar pattern of lower ES and OS. Unaffected FDRs of SCZ had lower ES, while those of BD showed only a trend towards lower ES and OS. Diagnosis of OCD was associated with significantly higher NA and OS, with lower EC and ES. Familial risk of OCD, on the other hand, was associated only with higher NA and lower EC. Diagnosis of AUD was associated with higher NA, lower EC and higher OS. However, the familial risk of AUD was not significantly associated with any temperamental domains. **Supplementary Table 5** shows the associations in the affected individuals after adjusting for the effects of familial loading for other disorders. The results were mostly the same, except an additional finding of higher NA in BD.

**Figure 2.**
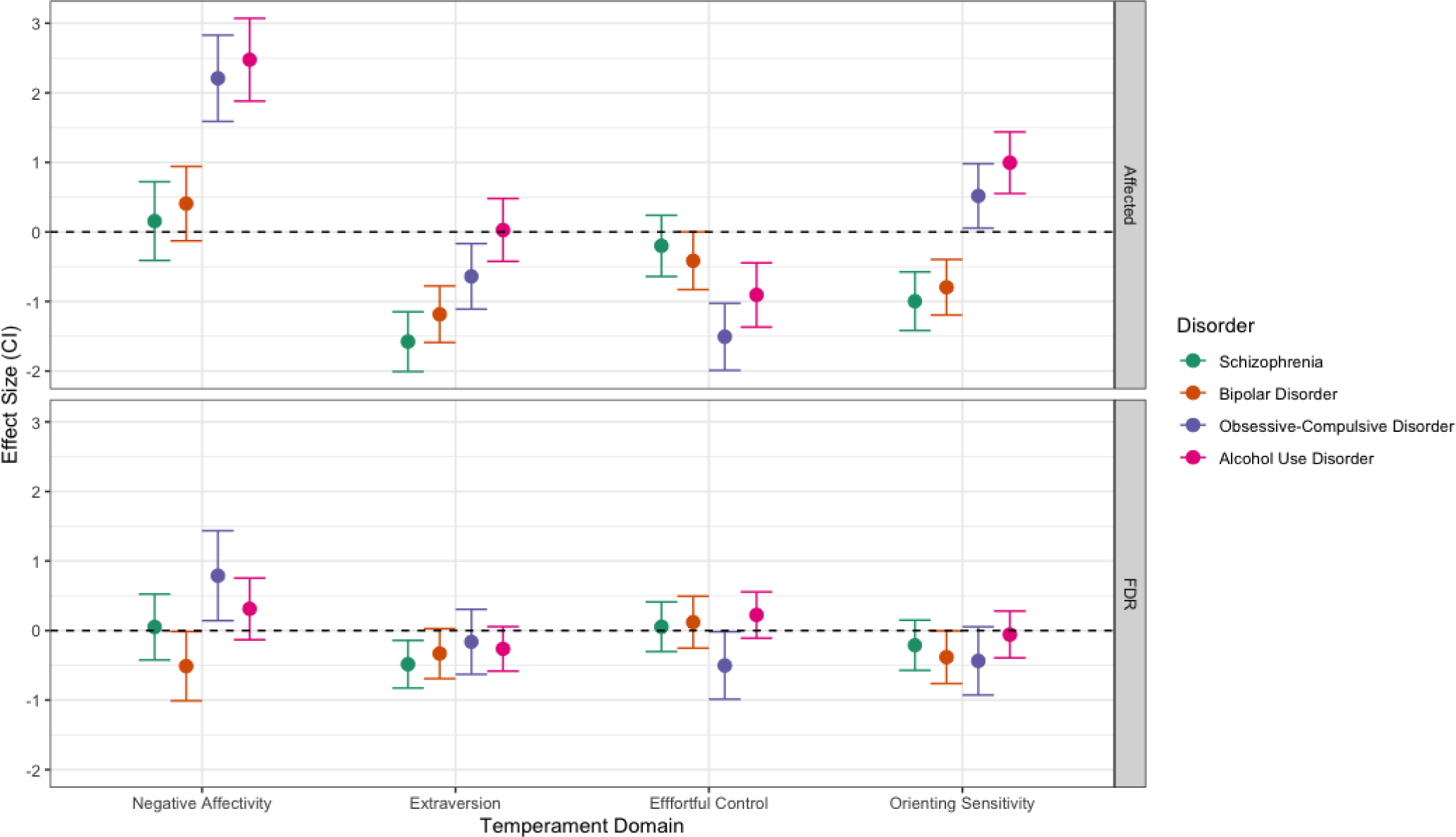
Effect sizes (Cohen’s d) of the comparison between each diagnostic group vs controls (upper panel), and familial risk vs controls (lower panel) FDR - First degree relative(s)

The results of the correlations between the temperamental domains with the illness severity (CGI-S) are shown in **Supplementary Figure 1**. EC was found to consistently correlate with CGI-S in all 4 disorders and NA with BD and OCD, while ES and OS with only AUD. Though statistically significant, these correlations were of weak/low magnitude (τ ranging 0.15-0.22).

The results of the multinomial Bayesian regression model, with temperament x ACEs interactions as predictors are shown in **Figure 4, Supplementary Table 6** and **Supplementary Figure 2**. Convergence of the MCMC models was found for all estimated parameters (*R̂* ≈ 1), and the cross-validation found a good fit in 96.1% of the leave-one-out samples. The Bayes factor for this model over the main-effects only model was 74.82, indicating “very strong” evidence (Lee & Wagenmakers, 2014; Quintana & Williams, 2018) in favor of our main ‘temperament x ACEs’ model. Looking at the mean posterior probabilities (**Figure 3)**, the main effect of ACEs was found significantly associated only with AUD, though OCD and to a lesser extent BD, showed a trend to higher ACEs. Similarly, the interaction between ACEs and temperamental domains was found only in AUD, with ES (positive association), and EC (negative association). **Supplementary Figure 3** shows the results of the external validation of our final model. We found that the model had excellent prediction accuracy for AUD (AUC=0.82), acceptable for BD (AUC=0.74), OCD (AUC=0.75). The performance for prediction schizophrenia, however, was chance-level (AUC = 0.52).

**Figure 3.**
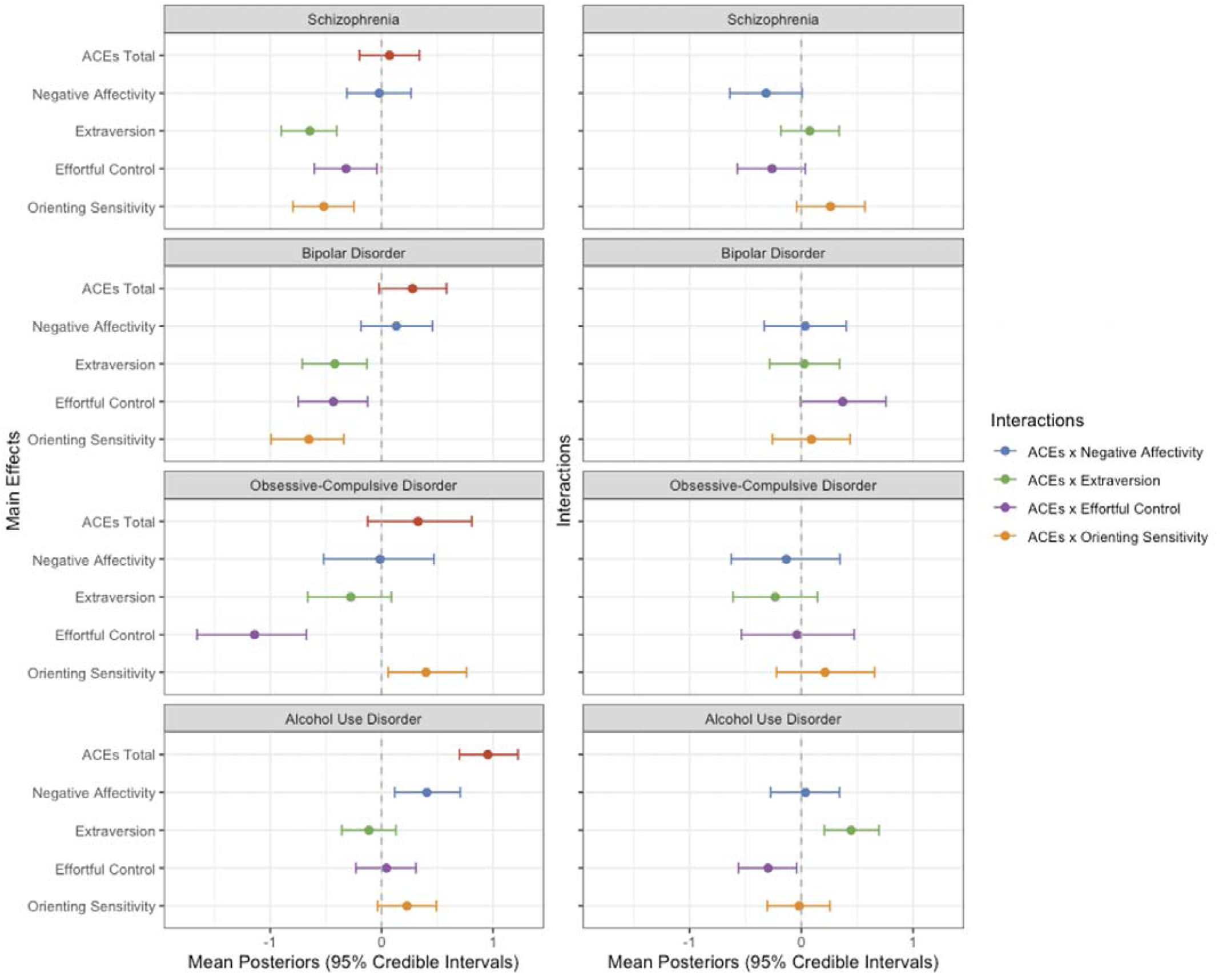
Results of the Bayesian multilevel regression models predicting diagnosis status by temperamental domains and adverse childhood experiences (ACEs), their main effects (left panel) and their interactions (right panel), adjusting for intra-familial correlations.

**Figure 4.**
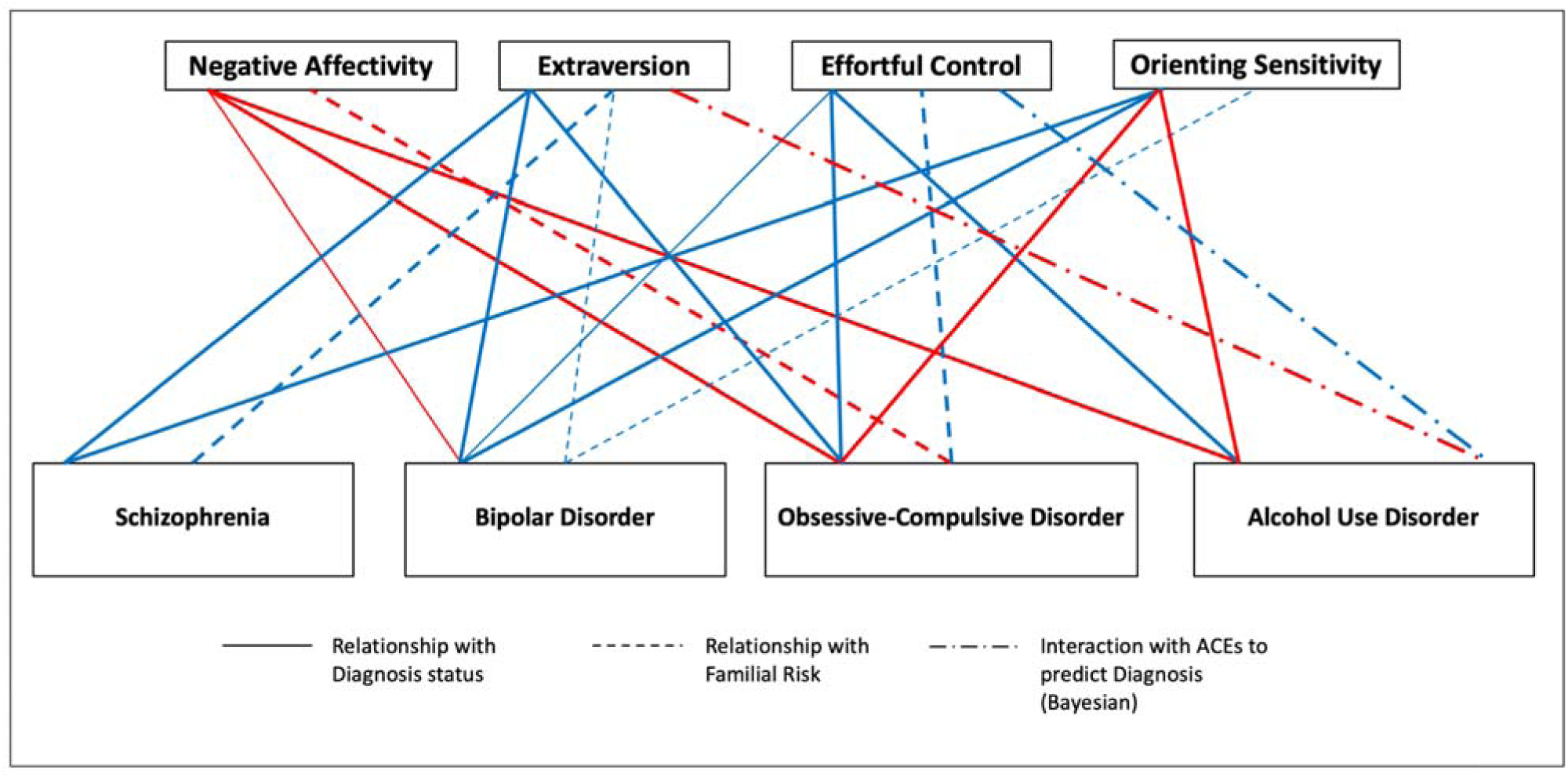
Summary of main results, showing significant associations between temperamental domains, their interactions with adverse childhood experiences (ACEs), with each diagnosis and familial risk. Red edges indicate positive associations, blue ones indicate negative association. Edges with smaller weights indicate significant findings that did not meet the Bonferroni threshold.

## Discussion

The current study explored various aspects of temperament in a large sample of adults from multiplex families of 4 major psychiatric disorders (SCZ, BD, OCD, AUD). This unique, transdiagnostic, genetically enriched and deeply phenotyped family-based sample enables us to comprehensively examine several questions about the endophenotypic nature of temperamental domains and their interaction with ACEs. We found many associations between each diagnosis, familial risk and temperamental domains.

First, we demonstrate that temperamental profiles are shared within families. The heritability of temperamental profiles, from twin studies has been found to be between 30-60% (Cloninger & Zwir, 2018; Gillespie et al., 2003). A genome-wide association study (GWAS) (Zwir et al, 2018) of temperament in healthy participants found the heritability explained by single nucleotide polymorphisms (h^2^-SNP) was also close to the same range (37-53%). This is substantially higher than the h^2^-SNP found in most psychiatric disorders (Mahjani et al., 2021; Mullins et al., 2021; Trubetskoy et al., 2022; Zhou et al., 2020), indicating that common genetic variants that are tested in GWAS, possibly have a greater, possibly more direct association with temperamental antecedents than with the syndromes themselves. In our study, considering the significant ‘within-family’ heterogeneity in terms of different diagnoses, there was still a significant degree of shared temperament.

Second, we show that temperamental profiles were associated with the 4 major disorders in several ways: in affected individuals (with modest correlations to illness severity), and their unaffected FDRs. Even after correction for multiple comparisons, many significant associations were found. We also modeled how some of these relationships may be moderated by the effects of childhood adversity, and found such associations only with AUD. In order to disentangle the numerous associations and interactions, we summarize these results in **Figure 4**, and discuss these relationships by each temperamental domain.

### Extraversion/Surgency domain

This domain is related to positive emotionality, pleasure/novelty seeking and higher activity levels, and is closely linked to the reward circuitry with dopamine as a key neurotransmitter (Wacker & Smillie, 2015). We found negative associations with the diagnoses of SCZ and BD, and with the familial risk of SCZ. Lower extraversion is one key dimension in schizotypy (Everett & Linscott, 2015; Grant et al., 2018), and is reported in many studies of SCZ and FDRs. Temperament findings in BD are found to be highly variable and state-prone (Barnett et al., 2011), most studies in euthymic BD report higher reward dependence/novelty seeking in the TCI (Zaninotto et al., 2016). However, we also note that many of those with BD diagnosis in our cohort had psychotic mood episodes (Sreeraj et al., 2021), plus we dummy-coded ‘schizoaffective disorder’ as both SCZ and BD. This may have led to the overlap in our findings, or indicate extraversion as a transdiagnostic endophenotype for psychosis.

Interaction of extraversion and ACEs was positively associated with AUD. It is interesting to note, that the main/direct association extraversion was not significant (even without including ACEs as a predictor), while only the interaction was. Most studies in AUD have been relatively consistent that such interactions have a much greater role, than either in isolation (Dick & Kendler, 2012; Lukasiewicz et al., 2008).

### Negative Affectivity domain

This measures the tendency to experience a broad range of negative emotions (fear, frustration, sadness, discomfort), and correlates strongly with the harm avoidance domain of the TCI, and the neuroticism construct in personality assessments. Responses to acute threat (fear), potential harm (anxiety), and loss (sadness) are considered by the RDoC to be central constructs within the negative valence system (Hasratian et al., 2022). They have been most commonly studied in the context of depressive and anxiety disorders. In our study, we found strong associations among those with a diagnosis of OCD and AUD, and also BD (after adjusting for familial risk).

### Effortful control domain

Effortful control is a key domain that determines self-regulation and is largely based upon the functioning of the frontal executive attention networks. It is conceptualized as the capacity to override impulsive, reactive responses and substitute responses in service of long-term goals (Eisenberg et al., 2004). Thus, it reflects the capacity for executive control of cognition and behavior, including the ability to focus, shift, and sustain attention (i.e., attention control), inhibit inappropriate responses (i.e., inhibitory control), and initiate approach responses despite reactive motivation to avoid or a lack of approach motivation (i.e., activation control).

We found lower effortful control in those with a diagnosis of OCD and AUD, and trend-level in BD. A large amount of existing literature supports these findings. Lower effortful control was found associated with familial risk of OCD, which is again supported by existing neurocognitive studies of response inhibition deficits in unaffected FDRs of OCD (de Wit et al., 2012; Menzies et al., 2007). This configuration with lower effortful control along with higher negative affectivity seen associated with both diagnosis as well as familial risk of OCD may well be an endophenotypic profile of temperament in OCD. We also see a highly significant interaction with ACEs in AUD, showing a negative association, which is in an opposite direction from that of the extraversion/surgency domain. This might be a profile of temperamental x environmental risk unique to AUD.

### Orienting sensitivity domain

This domain taps into the “posterior attention network”, which is the ability to prioritize sensory input by selecting a modality or location, and is modulated by the cholinergic systems in the brain (Petersen & Posner, 2012). We found significant associations with all 4 diagnoses: negative in SCZ and BD, positive in OCD and AUD. No differences were found between the FDRs for any disorder and healthy controls. There is a wealth of research on attentional impairments in SCZ and BD (Bora & Murray, 2014; Huang et al., 2019). Some research indicates that deficits in ability to filter out “cues” are strongly correlated to psychosis, particularly positive psychotic symptoms in both the disorders (Frydecka et al., 2014; Morris et al., 2013). However, no associations between familial risk of any of the 4 disorders was found with this domain, raising the question of whether this may be more of an illness/state-dependent marker rather than an endophenotype.

### Summary and Implications

We find that extraversion is a transdiagnostic temperamental dimension, being low in SZ, BD and OCD and high in AUD through its interactions with ACE. Orienting sensitivity is another transdiagnostic temperamental dimension being low in SCZ/BD and high in OCD/AUD. These findings within affected individuals are largely consistent with those of many smaller studies that have examined temperament within each psychiatric disorder separately. The temperamental factors associated with familial risk of SCZ (lower extraversion) and OCD (higher negative affectivity, lower effortful control) among FDRs also support previous research, while there were no specific traits in unaffected individuals associated with familial risk of BD and AUD. The influence of ACEs, as a main effect or as an interaction with temperamental domains, was noted only in AUD. Our study offers useful insights, as to how these common temperamental factors, with various nuanced configurations, play out in these psychiatric disorders.

### Strengths

To our knowledge, ours is the first study to examine temperament trans-diagnostically across a large sample of adults. A key, unique aspect of our sample is the systematic evaluation of persons from a genetically-enriched cohort of multiplex families. We were also able to demonstrate the reliability of our temperament assessments, across the considerable language diversity within our sample from India. Third, we used several robust statistical approaches, both frequentist and bayesian, to statistically model these complex relationships between multiple disorders, their familial risk, temperamental profiles and childhood adversity. Last, though our main discovery sample comprised multiplex (or “genetically-enriched”) families, we could demonstrate that our findings may be generalizable to other simplex/sporadic cases by testing the external validity in an independent sample. Our approach to predictive modeling also conforms to the best practices advocated for psychiatry from a recent systematic review (Meehan et al., 2022).

### Limitations

Although we work under the assumption that temperamental factors are likely to be “antecedents”, our study was cross-sectional in nature, and self-reported temperament in adulthood could be state-dependent or consequences of various illness-related factors. A prospective design with follow-up of the “at-risk” FDRs until conversion to disease status may provide the best evidence for the predictive value of these factors, and this would be within the scope of ongoing longitudinal follow-up phase of ADBS. We also have not analyzed interactions within specific temperamental domains (e.g negative affectivity * effortful control), or have not teased out the effects of various types of childhood adversities, which could help in further refining our risk characterization.

### Conclusions and Future Directions

Temperamental antecedents, by their very nature, may provide us with some of the best clinical predictors of risk of developing various psychiatric disorders, owing to their measurability at any age group, their stability across time, along with strong interactions with environmental factors. Integrating temperamental domains could improve our precision in definitions/criteria of “high risk” or “ultra high risk” groups for psychiatric disorders, beyond family history and before the appearance of sub-syndromal symptomatology. This could prove to be crucial for early intervention. As increasing evidence maps temperamental domains more clearly to neural circuits and genetic factors, this might offer further translational leads to evolve better preventive or treatment strategies for psychiatric disorders.

## Acknowledgments

Nil

## Data availability statement

The study datasets will be made available upon request to the Principal Investigators of the ongoing ADBS (adbsnimhans.org) and c-VEDA (cveda-project.org) studies. The R codes used for statistical analysis can be provided upon request to the corresponding author (SB).

---

**Supplementary Table 1.** Internal consistency (Cronbach’s alpha) of each domain and sub-domain within the self-reported adult temperament questionnaire (ATQ) used in the study for each language.

| Factor | Sub-factor | Definition | Cronbach's alpha |  |  |  |  |  |  |
| --- | --- | --- | --- | --- | --- | --- | --- | --- | --- |
|  |  |  | English | Hindi | Kannada | Telugu | Tamil | Malayalam | Overall |
| Negative Affectivity | Fear (7) | Unpleasant affect associated with the anticipation of pain or distress. | 0.76 | 0.77 | 0.74 | 0.71 | 0.65 | 0.70 | 0.76 |
|  | Frustration (6) | The amount of unpleasant affect related to the interruption of ongoing tasks and behavior or to the blocking of a desired goal. | 0.64 | 0.68 | 0.64 | 0.61 | 0.71 | 0.71 | 0.64 |
|  | Sadness (7) | The amount of lowered mood that is related to object or person loss, disappointment, and exposure to suffering. | 0.65 | 0.72 | 0.66 | 0.60 | 0.70 | 0.65 | 0.65 |
|  | Discomfort (6) | The amount of unpleasant affect resulting from the sensory qualities of stimulation, including irritation, pain, and discomfort resulting in the intensity, rate, complexity of light, movement sound, smell/taste, temperature, and texture. | 0.82 | 0.87 | 0.82 | 0.79 | 0.71 | 0.82 | 0.82 |
| Effortful control | Activation control (7) | The capacity to suppress negatively toned impulses and thereby resist the execution of inappropriate avoidance tendencies | 0.51 | 0.61 | 0.70 | 0.70 | 0.57 | 0.70 | 0.51 |
|  | Inhibitory control (7) | The capacity to suppress positively toned impulses and thereby resist the execution of inappropriate approach tendencies | 0.62 | 0.60 | 0.53 | 0.71 | 0.64 | 0.64 | 0.62 |
|  | Attentional control (5) | The ability to intentionally shift and focus attention in relation to task relevant stimuli and thoughts. | 0.58 | 0.63 | 0.67 | 0.57 | 0.57 | 0.61 | 0.58 |
| Extraversion | Sociability (5) | The pleasure about social interaction and the presence of others | 0.71 | 0.63 | 0.73 | 0.60 | 0.67 | 0.74 | 0.71 |
|  | High intensity Pleasure (7) | The pleasure about situations with high intensity stimulation, rate, complexity, novelty and incongruence | 0.70 | 0.61 | 0.68 | 0.70 | 0.67 | 0.67 | 0.70 |
|  | Positive affect (5) | Latency, threshold, intensity, duration and frequency for the experience of pleasure | 0.56 | 0.58 | 0.53 | 0.55 | 0.65 | 0.62 | 0.56 |
| Orienting sensitivity | Neutral perceptual sensitivity (5) | Notice stimulation with low intensity in the own body or in the environment | 0.68 | 0.64 | 0.69 | 0.62 | 0.74 | 0.73 | 0.68 |
|  | Affective perceptual sensitivity (5) | Spontaneous emotional and aware cognitions combined with low intensity stimulation | 0.70 | 0.66 | 0.72 | 0.67 | 0.58 | 0.68 | 0.70 |
|  | Associative sensitivity (5) | Spontaneous cognition without a direct association in the environment | 0.72 | 0.71 | 0.73 | 0.68 | 0.72 | 0.70 | 0.72 |

**Supplementary Table 2.** Raw scores of each adult temperamental domain and total adverse childhood experiences (ACEs) score in the full sample (N=1162), divided by diagnostic and familial risk group.

| ATQ Domain | Schizophrenia |  | Bipolar Disorder |  | Obsessive-Compulsive Disorder |  | Alcohol Use Disorder |  | Healthy Controls |
| --- | --- | --- | --- | --- | --- | --- | --- | --- | --- |
|  | Aff | FDRs | Aff | FDRs | Aff | FDRs | Aff | FDRs |  |
| Negative Affectivity | 15.21<br>(3.20) | 15.06<br>(2.85) | 15.43<br>(3.06) | 14.75<br>(2.84) | 17.18<br>(2.89) | 15.99<br>(2.23) | 16.72<br>(2.63) | 15.29<br>(2.70) | 14.55<br>(2.38) |
| Effortful Control | 13.00<br>(2.51) | 13.77<br>(2.00) | 12.82<br>(2.21) | 13.74<br>(2.19) | 11.70<br>(2.24) | 13.10<br>(2.07) | 12.62<br>(2.53) | 13.96<br>(1.99) | 13.56<br>(1.83) |
| Extraversion | 11.16<br>(2.29) | 12.13<br>(1.93) | 11.49<br>(2.09) | 12.27<br>(1.94) | 12.12<br>(2.19) | 12.47<br>(2.11) | 12.33<br>(2.42) | 12.49<br>(2.00) | 13.39<br>(1.85) |
| Orienting Sensitivity | 10.64<br>(2.42) | 11.71<br>(2.11) | 10.82<br>(2.30) | 11.69<br>(1.98) | 12.41<br>(2.02) | 11.88<br>(1.93) | 12.14<br>(2.02) | 11.92<br>(1.98) | 12.30<br>(2.04) |
| ACEs Total Score | 8.83<br>(7.72) | 8.87<br>(7.36) | 9.29<br>(7.07) | 7.92<br>(6.76) | 9.86<br>(6.65) | 7.76<br>(6.56) | 16.12<br>(9.29) | 9.91<br>(7.99) | 6.67 (6.24) |

**Supplementary Table 3.** Relationship between adult temperamental domains with covariates - age, sex and education along with the intrafamilial concordance in the full sample (N=1162)

|  | Negative Affectivity |  | Effortful Control |  | Extraversion |  | Orienting Sensitivity |  |
| --- | --- | --- | --- | --- | --- | --- | --- | --- |
|  | B (SE) | p-value | B (SE) | p-value | B (SE) | p-value | B (SE) | p-value |
| Intercept | 14.276<br>(0.575) | <0.001 | 13.103<br>(0.437) | <0.001 | 14.163<br>(0.434) | <0.001 | 11.09<br>(0.441) | <0.001 |
| Age (in years) | 0.012<br>(0.008) | 0.154 | 0.021<br>(0.006) | 0.001* | -0.037<br>(0.006) | <0.001* | 0.004<br>(0.006) | 0.526 |
| Gender (1=Male;<br>0=Female) | -0.83<br>(0.226) | <0.001* | -0.437<br>(0.171) | 0.011* | 0.059<br>(0.17) | 0.728 | -0.356<br>(0.173) | 0.04* |
| Education (in years) | 0.051<br>(0.027) | 0.057 | -0.003<br>(0.02) | 0.865 | 0.011<br>(0.02) | 0.6 | 0.097<br>(0.021) | <0.001* |
| Intra-familial concordance<br>(ICC & 95% CIs) | 0.271<br>(0.199 - 0.341) |  | 0.091<br>(0.025 - 0.157) |  | 0.194<br>(0.124 - 0.259) |  | 0.259<br>(0.189 - 0.330) |  |

ICC - Intraclass correlation coefficient; CI - Confidence Intervals; SE = Standard Error

**Supplementary Table 4.** Association between each diagnosis or familial risk group with temperamental domains, by linear mixed effects regression with age, gender and education as fixed-effects covariates, family ID as random effects covariate.

| ATQ Domain | Schizophrenia |  | Bipolar Disorder |  | Obsessive-Compulsive Disorder |  | Alcohol Use Disorder |  |
| --- | --- | --- | --- | --- | --- | --- | --- | --- |
|  | B (SE) | p-value | B (SE) | p-value | B (SE) | p-value | B (SE) | p-value |
| **Within affected individuals (n=523) vs healthy controls (n=173)** |  |  |  |  |  |  |  |  |
| Negative Affectivity | 0.016<br>(0.288) | 0.957 | 0.449<br>(0.277) | 0.105 | **2.314**<br>**(0.319)** | **<0.001** | **2.456**<br>**(0.307)** | **<0.001** |
| Effortful Control | -0.202<br>(0.228) | 0.378 | -0.503<br>(0.219) | 0.022 | **-1.57**<br>**(0.252)** | **<0.001** | **-0.83**<br>**(0.243)** | **0.001** |
| Extraversion | **-1.529**<br>**(0.221)** | **<0.001** | **-1.234**<br>**(0.212)** | **<0.001** | **-0.62**<br>**(0.245)** | **0.011** | -0.111<br>(0.236) | 0.639 |
| Orienting Sensitivity | **-1.039**<br>**(0.214)** | **<0.001** | **-0.828**<br>**(0.206)** | **<0.001** | **0.607**<br>**(0.237)** | **0.011** | **0.929**<br>**(0.229)** | **<0.001** |
| **Within unaffected individuals with familial risk (N=457) vs healthy controls (n=173)** |  |  |  |  |  |  |  |  |
| Negative Affectivity | -0.036<br>(0.244) | 0.881 | -0.439<br>(0.254) | 0.085 | **0.983**<br>**(0.336)** | **0.004** | 0.335<br>(0.226) | 0.139 |
| Effortful Control | -0.006<br>(0.183) | 0.975 | 0.06<br>(0.191) | 0.755 | **-0.587**<br>**(0.253)** | **0.02** | 0.269<br>(0.17) | 0.114 |
| Extraversion / Surgency | **-0.471**<br>**(0.179)** | **0.009** | -0.382<br>(0.187) | 0.042 | -0.107<br>(0.247) | 0.665 | -0.272<br>(0.166) | 0.103 |
| Orienting Sensitivity | -0.207<br>(0.185) | 0.263 | -0.402<br>(0.193) | 0.038 | -0.274<br>(0.255) | 0.284 | -0.004<br>(0.172) | 0.982 |

**Supplementary Table 5.** Associations between each diagnosis with the temperament domains after adjusting for the effect of familial risk.

| ATQ Domain | Schizophrenia |  | Bipolar Disorder |  | Obsessive-Compulsive Disorder |  | Alcohol Use Disorder |  |
| --- | --- | --- | --- | --- | --- | --- | --- | --- |
|  | B (SE) | p-value | B (SE) | p-value | B (SE) | p-value | B (SE) | p-value |
| Negative Affectivity | 0.416<br>(0.288) | 0.149 | **0.859**<br>**(0.273)** | **0.002** | **2.318**<br>**(0.316)** | **<0.001** | **2.235**<br>**(0.303)** | **<0.001** |
| Effortful Control | -0.259<br>(0.223) | 0.247 | -0.399<br>(0.212) | 0.06 | **-1.583**<br>**(0.245)** | **<0.001** | **-0.978**<br>**(0.235)** | **<0.001** |
| Extraversion / Surgency | **-0.884**<br>**(0.218)** | **<0.001** | **-0.608**<br>**(0.206)** | **0.003** | **-0.496**<br>**(0.239)** | **0.038** | -0.018<br>(0.229) | 0.938 |
| Orienting Sensitivity | -0.34<br>(0.213) | 0.11 | -0.021<br>(0.202) | 0.918 | **0.675**<br>**(0.233)** | **0.004** | **1.063**<br>**(0.224)** | **<0.001** |

**Supplementary Figure 1.**
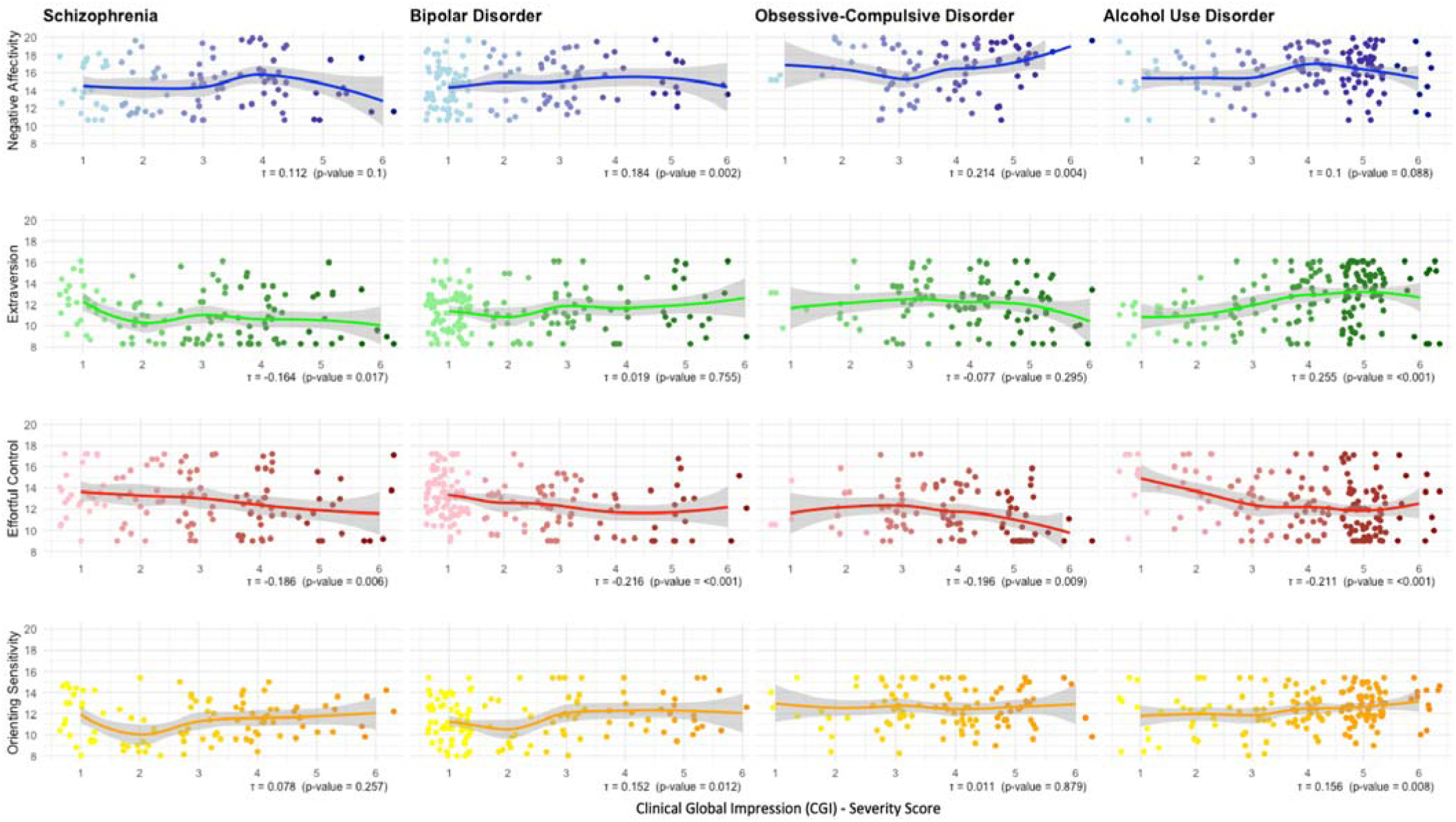
Correlations (Kendall’s tau) between current illness severity (Clinical Global Impression - Severity score) within each disorder and each temperamental domain. The x-axes (CGI-S scores) are on an ordinal/ likert scale between 1 to 7, but they have been jittered to reduce overlap and improve representation. The lines represent the “loess” (locally weighted scatterplot smoothing) regression lines

**Supplementary Table 6.** Results of the bayesian multinomial logistic regression looking for associations between each diagnostic category and temperament domain (after adjusting for the effects of age, gender, education and random intercepts for family ID)

| Outcome (Diagnosis) | Predictor | Estimate | l-95% CI | u-95% CI | R-hat | Bulk_ESS | Tail_ESS |
| --- | --- | --- | --- | --- | --- | --- | --- |
| Schizophrenia | Intercept | -2.917 | -3.43 | -2.488 | 1.003 | 2936 | 4640 |
| Bipolar Disorder | Intercept | -3.465 | -4.195 | -2.864 | 1.003 | 2768 | 4137 |
| Obsessive-Compulsive Disorder | Intercept | -4.947 | -6.303 | -3.907 | 1.001 | 2214 | 3776 |
| Alcohol Use Disorder | Intercept | -2.43 | -2.809 | -2.099 | 1.003 | 2198 | 4403 |
| Schizophrenia | ACEs Total | 0.071 | -0.199 | 0.339 | 1 | 11819 | 6345 |
| Schizophrenia | Negative Affectivity | -0.022 | -0.312 | 0.265 | 1 | 8387 | 6722 |
| Schizophrenia | Effortful Control | -0.319 | -0.604 | -0.043 | 1.002 | 8929 | 5921 |
| Schizophrenia | Extraversion | -0.644 | -0.901 | -0.403 | 1 | 9835 | 6356 |
| Schizophrenia | Orienting Sensitivity | -0.519 | -0.795 | -0.249 | 1.001 | 8792 | 6852 |
| Schizophrenia | ACEs x Negative Affectivity | -0.315 | -0.64 | 0.006 | 1.001 | 7383 | 6402 |
| Schizophrenia | ACEs x Effortful Control | -0.262 | -0.572 | 0.036 | 1.001 | 8005 | 6376 |
| Schizophrenia | ACEs x Extraversion | 0.076 | -0.183 | 0.339 | 1.001 | 11624 | 6144 |
| Schizophrenia | ACEs x Orienting Sensitivity | 0.261 | -0.042 | 0.57 | 1.002 | 9955 | 6657 |
| Bipolar Disorder | ACEs Total | 0.277 | -0.023 | 0.582 | 1.001 | 9200 | 5767 |
| Bipolar Disorder | Negative Affectivity | 0.132 | -0.186 | 0.456 | 1.001 | 7390 | 6479 |
| Bipolar Disorder | Effortful Control | -0.433 | -0.748 | -0.126 | 1 | 8952 | 6412 |
| Bipolar Disorder | Extraversion | -0.42 | -0.712 | -0.132 | 1 | 8602 | 6533 |
| Bipolar Disorder | Orienting Sensitivity | -0.653 | -0.993 | -0.34 | 1 | 7128 | 6396 |
| Bipolar Disorder | ACEs x Negative Affectivity | 0.035 | -0.332 | 0.402 | 1.001 | 7580 | 6595 |
| Bipolar Disorder | ACEs x Effortful Control | 0.371 | -0.008 | 0.757 | 1 | 6409 | 6876 |
| Bipolar Disorder | ACEs x Extraversion | 0.026 | -0.284 | 0.343 | 1 | 9548 | 5967 |
| Bipolar Disorder | ACEs x Orienting Sensitivity | 0.091 | -0.259 | 0.436 | 1.001 | 8154 | 6366 |
| Obsessive-Compulsive Disorder | ACEs Total | 0.327 | -0.125 | 0.809 | 1 | 6781 | 6284 |
| Obsessive-Compulsive Disorder | Negative Affectivity | -0.014 | -0.52 | 0.469 | 1.001 | 4475 | 5422 |
| Obsessive-Compulsive Disorder | Effortful Control | -1.14 | -1.656 | -0.675 | 1 | 4915 | 5587 |
| Obsessive-Compulsive Disorder | Extraversion | -0.277 | -0.663 | 0.087 | 1.001 | 7405 | 6197 |
| Obsessive-Compulsive Disorder | Orienting Sensitivity | 0.398 | 0.059 | 0.762 | 1.001 | 5328 | 6126 |
| Obsessive-Compulsive Disorder | ACEs x Negative Affectivity | -0.134 | -0.627 | 0.346 | 1.001 | 7505 | 6496 |
| Obsessive-Compulsive Disorder | ACEs x Effortful Control | -0.038 | -0.535 | 0.473 | 1 | 7059 | 6274 |
| Obsessive-Compulsive Disorder | ACEs x Extraversion | -0.234 | -0.611 | 0.145 | 1.002 | 8556 | 6227 |
| Obsessive-Compulsive Disorder | ACEs x Orienting Sensitivity | 0.213 | -0.222 | 0.655 | 1.001 | 7528 | 6751 |
| Alcohol Use Disorder | ACEs Total | 0.953 | 0.699 | 1.224 | 1.001 | 6241 | 5916 |
| Alcohol Use Disorder | Negative Affectivity | 0.406 | 0.117 | 0.706 | 1 | 7871 | 6826 |
| Alcohol Use Disorder | Effortful Control | 0.043 | -0.23 | 0.307 | 1.001 | 8029 | 5236 |
| Alcohol Use Disorder | Extraversion | -0.114 | -0.357 | 0.129 | 1 | 7999 | 5962 |
| Alcohol Use Disorder | Orienting Sensitivity | 0.228 | -0.036 | 0.492 | 1 | 8746 | 6629 |
| Alcohol Use Disorder | ACEs x Negative Affectivity | 0.037 | -0.275 | 0.342 | 1.001 | 8189 | 6042 |
| Alcohol Use Disorder | ACEs x Effortful Control | -0.298 | -0.562 | -0.042 | 1.001 | 8124 | 6470 |
| Alcohol Use Disorder | ACEs x Extraversion | 0.446 | 0.206 | 0.695 | 1.001 | 8133 | 5762 |
| Alcohol Use Disorder | ACEs x Orienting Sensitivity | -0.019 | -0.304 | 0.256 | 1.002 | 8501 | 6520 |

*CI - Credible Intervals; ESS - Effective sample size; ACEs - Adverse Childhood Experiences Total Score*

**Supplementary Figure 2.**
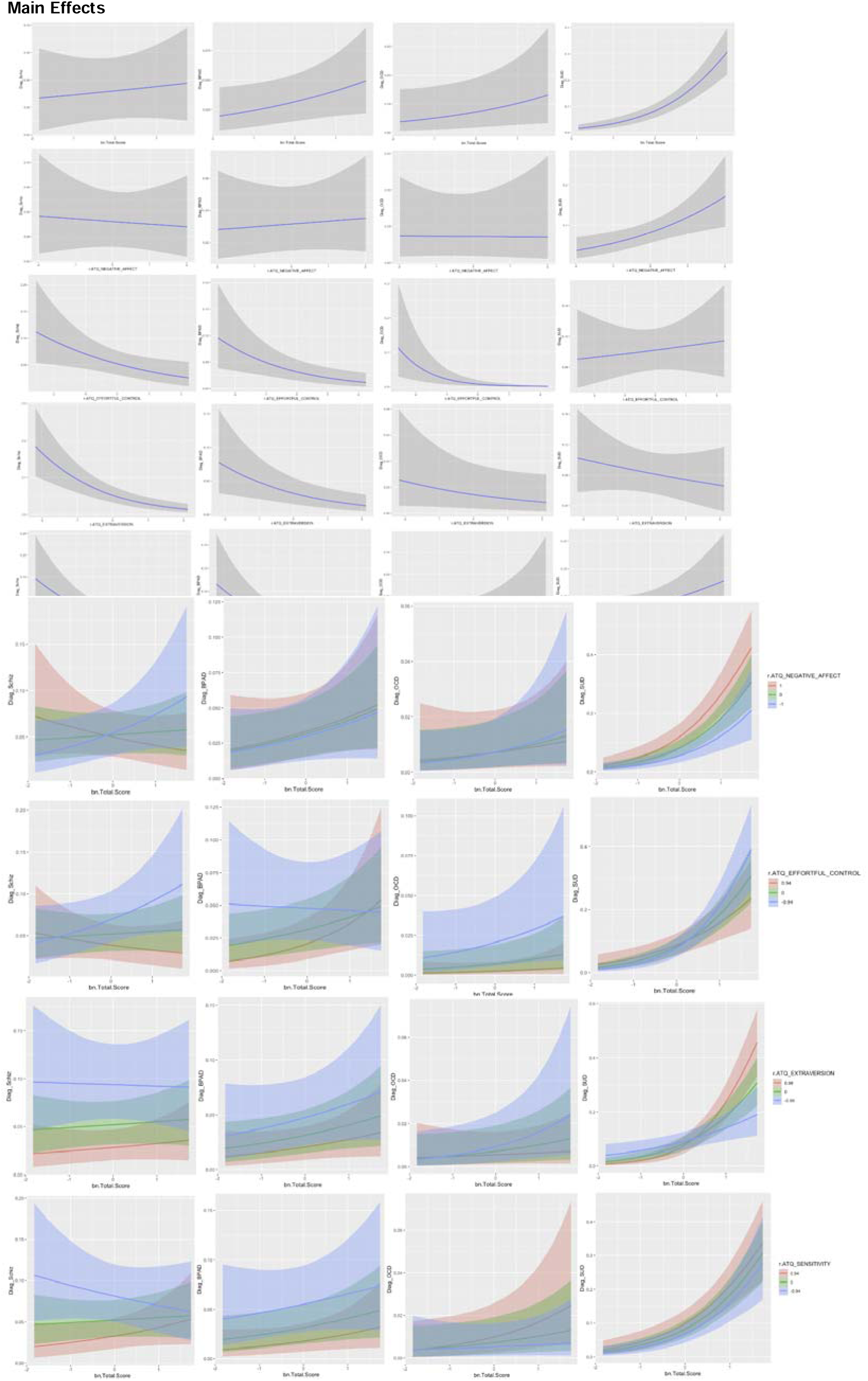
Conditional/marginal plots of the main effects and interactions between temperamental domains and childhood adversity predicting each diagnosis.

**Supplementary Table 7.** Characteristics of the validation dataset - adults (aged 18-23 years) in the Consortium for Vulnerability to Externalizing Disorders and Addictions (c-VEDA)

| Characteristic | Mean (SD) or N(%) |
| --- | --- |
| n | 2485 |
| Age | 19.84 (3.21) |
| Gender (male) | 998 (40.2%) |
| Years of Education | 13.46 (2.26) |
| ACEs Total Score | 7.12 (7.59) |
| ATQ Domains |  |
| Negative Affectivity | 16.86 (2.54) |
| Effortful Control | 12.26 (1.73) |
| Extraversion | 13.57 (1.89) |
| Orienting Sensitivity | 13.66 (2.12) |
| Psychiatric Diagnosis |  |
| Schizophrenia | 32 (1.3%) |
| Bipolar Disorder | 18 (0.7%) |
| Obsessive-Compulsive Disorder | 13 (0.5%) |
| Alcohol Use Disorder | 16 (0.6%) |

**Supplementary Figure 3.**
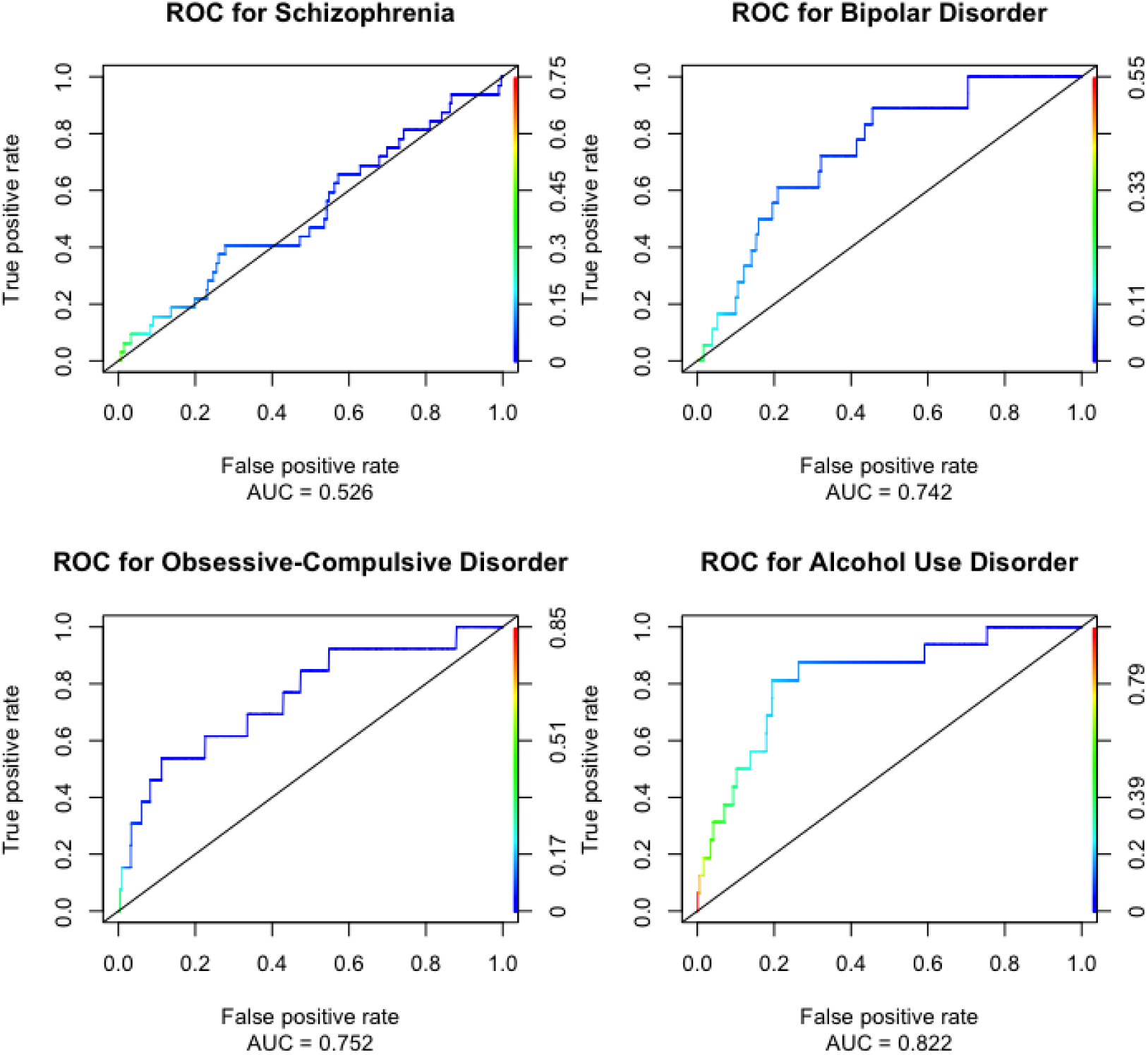
Receiver Operator Characteristic (ROC) plots for prediction of the final model in the cVEDA dataset.

